# Topological Feature Fusion for Dermoscopic Skin Cancer Detection

**DOI:** 10.1101/2025.11.25.25340992

**Authors:** Fulya Tastan, Sayoni Chakraborty, Sangyeon Lee, Baris Coskunuzer

## Abstract

Skin cancer is a common and potentially fatal disease where early detection can save lives, especially for melanoma. Current deep learning systems classify skin lesions well, but they mainly rely on appearance cues and may miss deeper structural patterns in lesions. We present TopoCon MP, a method that extracts multiparameter topological signatures from dermoscopic images to capture multiscale lesion structure, and fuses these signatures with Vision Transformers using a supervised contrastive objective. Across three public datasets, TopoCon MP consistently improves performance over strong pretrained CNN and ViT baselines, including in cross dataset transfer. Ablations show that multiparameter topology and contrastive fusion each contribute to the gains. The resulting topological channels also provide an interpretable view of lesion organization, aligning with clinically meaningful structures. Overall, TopoCon MP demonstrates that multipersistence based topology can serve as a complementary modality for robust skin cancer detection.

## 1. Introduction

Skin cancer is among the most common and potentially lethal malignancies worldwide, so early and accurate detection is essential for reducing morbidity and mortality. Clinical decision support systems based on machine learning have shown strong promise for automating the analysis of dermoscopic images, with deep models approaching dermatologist-level performance on lesion classification tasks (Codella et al., 2018; Tang et al., 2020). However, conventional CNN and transformer architectures are driven primarily by local pixel and texture cues and may underutilize global properties such as lesion shape, connectivity patterns, and boundary irregularities that often distinguish malignant from benign lesions (Gutman et al., 2016).

Topological data analysis (TDA) offers a complementary perspective by extracting stable, multiscale descriptors of an image’s geometric and connectivity structure (Carlsson, 2009). Persistent homology summarizes how features such as connected components and holes appear and disappear across a filtration, yielding signatures that are robust to small deformations and illumination changes. Prior work has demonstrated the value of TDA in medical imaging, including tumor characterization and histopathology (Hofer et al., 2017), but almost all existing approaches rely on single-parameter filtrations and therefore cannot directly capture how multiple imaging cues interact across scales.

In this paper, we introduce *cubical multiparameter persistence* for dermoscopic image analysis and, to our knowledge, provide the first systematic study of multipersistence for skin cancer detection. We build coupled cubical filtrations that jointly encode intensity, scale, and simple texture surrogates, producing multiparameter Betti summaries that reflect cross-parameter interactions that single-parameter pipelines can miss. To fuse these topological signals with modern vision backbones, we integrate the multipersistence descriptors with a Vision Transformer (ViT) using *supervised contrastive learning*, encouraging class-consistent agreement between image features and topological structure in a shared representation space.

We evaluate our method, TopoCon-MP, on three publicly available skin lesion datasets that include melanoma, basal cell carcinoma, squamous cell carcinoma, and benign nevi. Experiments show that multipersistence alone is competitive with strong pretrained baselines, and that topology-aware contrastive fusion improves AUC, accuracy, and balanced accuracy over both CNN and ViT models, with particularly clear gains in cross-dataset transfer. In addition, the learned topological channels provide interpretable cues that co-localize with clinically meaningful structures such as pigment networks and irregular lesion borders.

Our contributions are:

- We present the first application and systematic evaluation of *cubical multiparameter persistence* for automated skin lesion classification.
- We propose a topology and vision fusion strategy that aligns multipersistence descriptors with a ViT using supervised contrastive learning to obtain joint representations.
- We demonstrate consistent performance gains over strong pretrained CNN and ViT baselines across multiple datasets, including a cross-dataset transfer setting, with ablations showing the advantage of multipersistence over single-parameter cubical persistence.
- We provide qualitative evidence that topological signals highlight clinically relevant morphology, supporting interpretability for dermatological imaging.

## 2. Background

### 2.1. Related Work

#### Machine learning methods in skin cancer detection

Machine learning, in particular deep learning, has transformed automated skin lesion analysis. Esteva et al. showed that convolutional neural networks (CNNs) trained on large collections of clinical and dermoscopic images can reach dermatologist-level performance on lesion classification (Esteva et al., 2017). Follow-up work fine-tuned ImageNet-pretrained CNNs such as ResNet on curated dermoscopy datasets (Menegola et al., 2017), examined robustness across cohorts (Brinker et al., 2019), and introduced benchmarks like HAM10000 that enabled large-scale evaluation and ensemble methods (Tschandl et al., 2018; Codella et al., 2018). More recent studies have explored multimodal models that fuse dermoscopy with patient metadata for improved risk stratification (Li and Shen, 2020) and transformer-based architectures that capture long-range spatial dependencies (Yuan et al., 2021). Despite this progress, challenges remain around data heterogeneity, interpretability, and deployment in real clinics (Patel et al., 2021). In particular, most models focus on pixel and texture cues and underutilize global properties such as lesion shape, connectivity, and boundary irregularity that are central to dermoscopic diagnosis (Gutman et al., 2016).

#### Topological machine learning in medical image analysis

Topological data analysis (TDA) provides stable, multiscale descriptors of geometric and connectivity structure (Carlsson, 2009). Persistent homology (PH) has been applied in many biomedical settings, including modeling cell development (McGuirl et al., 2020), delineating tumor margins (Qaiser et al., 2019), analyzing brain connectivity (Saggar et al., 2018), and extracting genomic signatures (Lum et al., 2013); see Skaf et al. (Skaf and Laubenbacher, 2022) for a survey. Building on these ideas, topological deep learning integrates PH summaries into trainable models (Hofer et al., 2017; Adams et al., 2017), with reported gains in segmentation (Kahle et al., 2021; Santhirasekaram et al., 2023) and classification (Chachólski et al., 2019; Johnson et al., 2022). Applications to melanoma and skin lesion analysis have begun to appear (Maurya et al., 2024; Chung et al., 2018), but they typically use single-parameter filtrations and do not model interactions between multiple imaging cues.

Multiparameter persistent homology generalizes PH to filtrations indexed by more than one parameter and has been developed theoretically and algorithmically in recent years (Botnan and Lesnick, 2022; Loiseaux et al., 2023; Korkmaz et al., 2025). To our knowledge, it has not yet been explored for dermoscopic image analysis. Our work introduces cubical multiparameter persistence for skin cancer detection and studies both standalone topological models and hybrid models that fuse multipersistence summaries with Vision Transformers. This fills a gap between existing TDA-based approaches, which rely on single-parameter pipelines, and mainstream deep learning methods, which largely ignore explicit topological structure.

### 2.2. Cubical Persistence

Persistent homology (PH) is a core tool in TDA for extracting multiscale structure from data such as point clouds, networks, and images (Dey and Wang, 2022). We focus on its image variant, *cubical persistence*. A brief overview follows; see (Coskunuzer and Akçora, 2024) for details. PH proceeds in three steps:

- **Filtration**: build a nested sequence of topological spaces.
- **Persistence diagrams**: record feature births and deaths across the sequence.
- **Vectorization**: map diagrams to fixed-length representations for machine learning.

#### Step 1. Constructing filtrations

In images, filtrations are typically cubical. Starting from a grayscale (or single color-channel) image 𝒳 ∈ ℝ*^r^*^×^*^s^* with pixel values *γ_ij_* and a sequence of thresholds *t*_1_ *<* · · · *< t_N_*, we form a sublevel sequence 𝒳_1_ ⊂ · · · ⊂ 𝒳*_N_* with

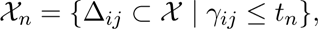

where Δ*_ij_* is the square pixel at position (*i, j*). Intuitively, pixels become “active” as the threshold increases, yielding a nested family of binary images (Fig. 1).

**Figure 1:**
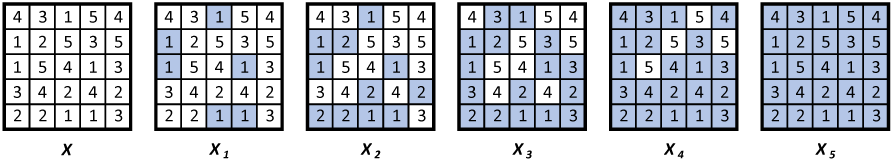
For the 5 × 5 image 𝒳 with the given pixel values, the sublevel filtration is the sequence of binary images 𝒳_1_ ⊂ · · · ⊂ 𝒳_5_.

#### Step 2. Persistence diagrams

PH tracks when topological features appear and dis-appear along {𝒳*_n_*}. If a feature *σ* is born at *t_m_* and disappears at *t_n_* with *m < n*, the pair (*b_σ_, d_σ_*) = (*t_m_, t_n_*) is added to the *k*-dimensional diagram PD*_k_*(𝒳), where *k* indexes connected components (*k* = 0), holes (*k* = 1), and higher-dimensional cavities. The lifespan *d_σ_* − *b_σ_* quantifies the prominence of the feature. In Fig. 1, PD_0_ captures connected components and PD_1_ captures holes.

#### Step 3. Vectorization

Since persistence diagrams are multisets of birth–death pairs, they are not directly suitable for standard learning architectures. Vectorization (Ali et al., 2023) converts diagrams into fixed-length representations. We primarily use *Betti vectors*, where *β_k_*(*t_n_*) counts the number of alive *k*-dimensional features at threshold *t_n_*, yielding 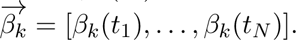 For example, for Fig. 1, 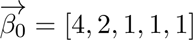 and 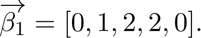 Alternative encodings include persistence images (Adams et al., 2017), landscapes (Bubenik and Dlotko, 2017), silhouettes (Chazal et al., 2014), and kernel methods (Ali et al., 2023). We favor Betti vectors for their efficiency, interpretability, and natural sequence form, which later extends to multiparameter Betti tensors and integrates well with transformer-based models.

## 3. Methodology

### 3.1. Cubical Multiparameter Persistence

#### From single to multiparameter persistence

In single-parameter cubical persistence, a grayscale (or single-channel) image 𝒳 induces a filtration 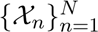 indexed by thresholds {*t_n_*}, where 𝒳*_n_* is the binary image obtained by activating pixels with intensity below *t_n_*. Persistent homology then tracks when connected components, holes, and higher-dimensional features appear and disappear as the threshold increases, and summarizes them with barcodes or persistence diagrams.

In multiparameter persistence, we let the image evolve along two or more directions at once. We focus on the two-parameter case. A *bifiltration* of an *r* × *s* image 𝒳 is a family {𝒳*_m,n_*} of binary images such that 𝒳*_m,n_* ⊂ 𝒳*_m_*_+1_*_,n_* and 𝒳*_m,n_* ⊂ 𝒳*_m,n_*_+1_. Each row and each column is a standard 1D filtration, and together they form a grid of nested images. Applying homology at each grid point gives a collection of topological features that now live over a 2D index set instead of a single line.

A key difference from the single-parameter case is that there is no unique way to assign a single birth and death time to each feature, since the indices (*m, n*) are only partially ordered. As a result, there is no canonical barcode or persistence diagram in general multiparameter settings (Botnan and Lesnick, 2022). Several alternative summaries have been proposed, such as rank invariants and multipersistence landscapes (Vipond et al., 2021; Loiseaux et al., 2023), but most of them are still relatively heavy for practical large-scale imaging.

#### Betti tensors as multiparameter signatures

In this work we adopt a simple but effective summary based on Betti numbers over the grid. For each grid point (*m, n*) and each homological dimension *k*, we define 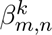 = number of *k*-dimensional topological features in 𝒳*_m,n_.* Collecting these values over the grid yields a 2D *Betti tensor* 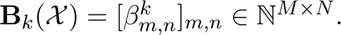

Intuitively, **B**_0_(𝒳) records how the number of connected components changes across two parameters, and **B**_1_(𝒳) does the same for holes. Unlike persistence diagrams, Betti tensors do not distinguish long-lived from short-lived features, but they provide a compact, grid-aligned representation that is easy to store and to feed into neural networks. This type of Betti-based encoding has been empirically effective in several medical and histopathological imaging tasks (Qaiser et al., 2019; Yadav et al., 2023; Du et al., 2022; Ali et al., 2023). Additional details and a more formal connection to the multipersistence literature are given in Appendix A.

#### Color multifiltrations for dermoscopic images

RGB dermoscopic images naturally support multiparameter filtrations (Korkmaz et al., 2025). Let 𝒳 be an RGB image with channel values *R_ij_, G_ij_, B_ij_* ∈ [0, 255] for each pixel (cubical cell) Δ*_ij_*. In general, choosing threshold sets 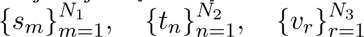 for the three channels defines a three-parameter multifiltration 𝒳*_m,n,r_* = {Δ*_ij_* ⊂ 𝒳 | *R_ij_* ≤ *s_m_, G_ij_* ≤ *t_n_, B_ij_* ≤ *v_r_*}, and corresponding 3D Betti tensors 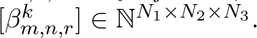 Figure 2 shows a small toy example with a 3 × 3 grid for clarity.

**Figure 2:**
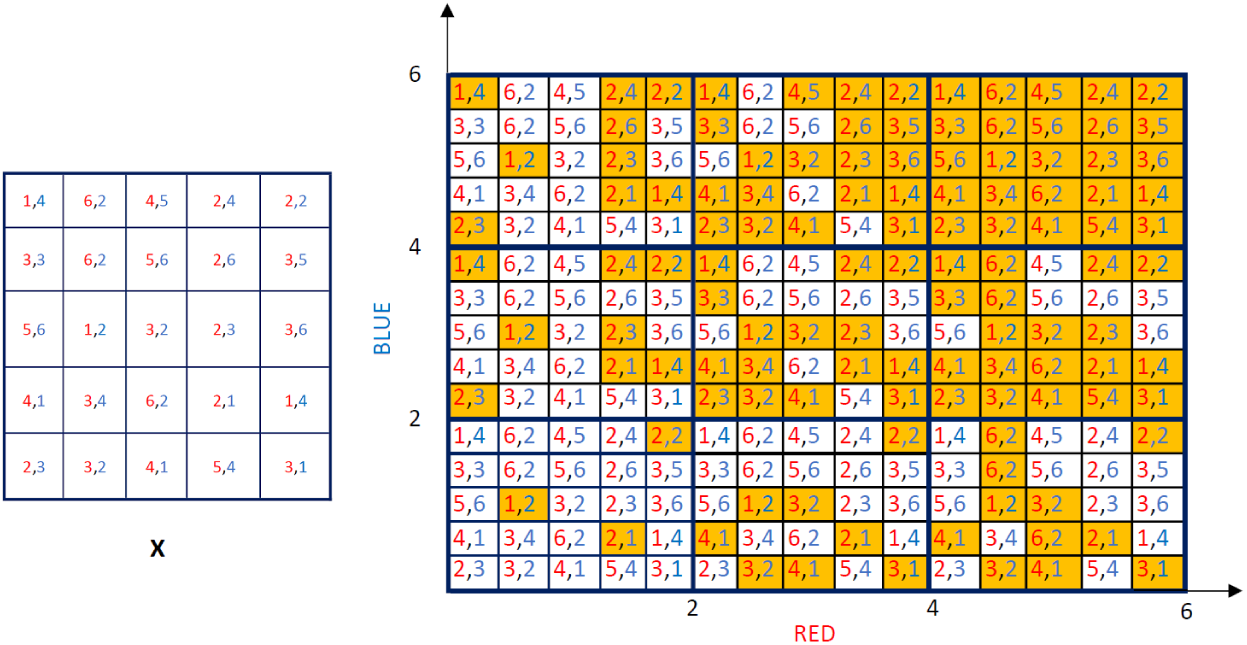
Toy example. For an image 𝒳 with two color channels, a simple color bifiltration produces a 3 × 3 grid of binary images (the actual grid we used is 20 × 20). Horizontally, pixels are activated (colored orange) when their red value falls below the threshold, and vertically, activation depends on the blue value. Each row and column forms an ordinary one dimensional filtration, while the grid as a whole defines a two dimensional multiparameter filtration.

For our experiments we adopt a computationally efficient two-parameter specialization tailored to dermoscopic images. We construct a bifiltration over the red and green channels (identified as most informative on a validation set) using *M* = *N* = 20 thresholds to form a 20 × 20 grid. For each image we compute the corresponding *β*_0_ and *β*_1_ tensors together with activated-pixel counts, and stack them into a 3 × 20 × 20 *topological image*. This multipersistence representation is used both as input to XGBoost baselines and as the topological branch in the TopoCon-MP fusion model.

**Figure 3:**
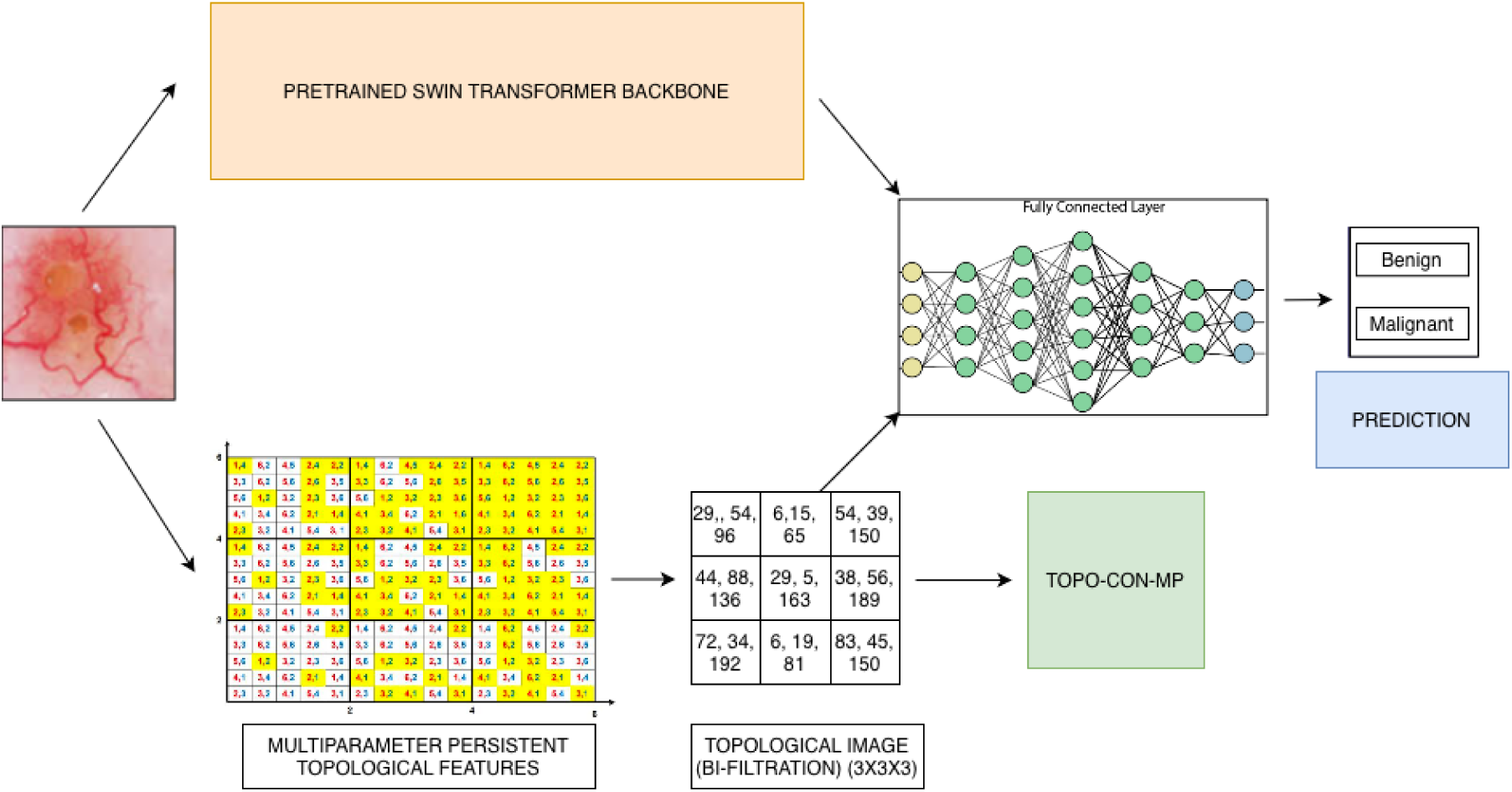
Overview of TopoCon-MP. The raw dermoscopic image is processed by a pretrained Swin Transformer backbone to obtain semantic image features. In parallel, we compute multiparameter Betti tensors on a fixed grid and stack *β*_0_, *β*_1_, and activated-pixel counts into a 3 × 20 × 20 topological image. This topological image is encoded with an MLP and aligned with the Swin features via a topology-aware supervised contrastive loss. The fused representation is passed to a final classifier for benign vs. malignant prediction.

A more formal discussion of multiparameter persistence, barcode obstructions, and alternative summaries (including our Betti tensor view) is provided in Appendix A.

### 3.2. Topology Aware Supervised Contrastive Learning

Supervised contrastive learning encourages representations that cluster samples from the same class while separating those from different classes. Standard supervised contrastive methods typically construct multiple “views” of each image using random augmentations such as cropping, rotation, or intensity jitter. In dermoscopy, however, aggressive spatial augmentations can distort lesion boundaries or alter diagnostically relevant texture patterns.

We therefore propose a *topology aware* supervised contrastive framework that uses the original dermoscopic image and its multiparameter topological embedding as two semantically consistent views of the same case. For each input image *I*, we compute its cubical multiparameter persistence representation, capturing structural and morphological characteristics in a label preserving and anatomically coherent way. The resulting bifiltration produces a 2D topological image Ψ(*I*) ∈ ℝ*^H^*^×^*^W^* ^×3^, where the three channels correspond to *β*_0_, *β*_1_, and the activated pixel map derived from the multipersistence computation. This RGB style topological image emphasizes global topology and boundary structure without introducing augmentation induced bias.

**Figure 4:**
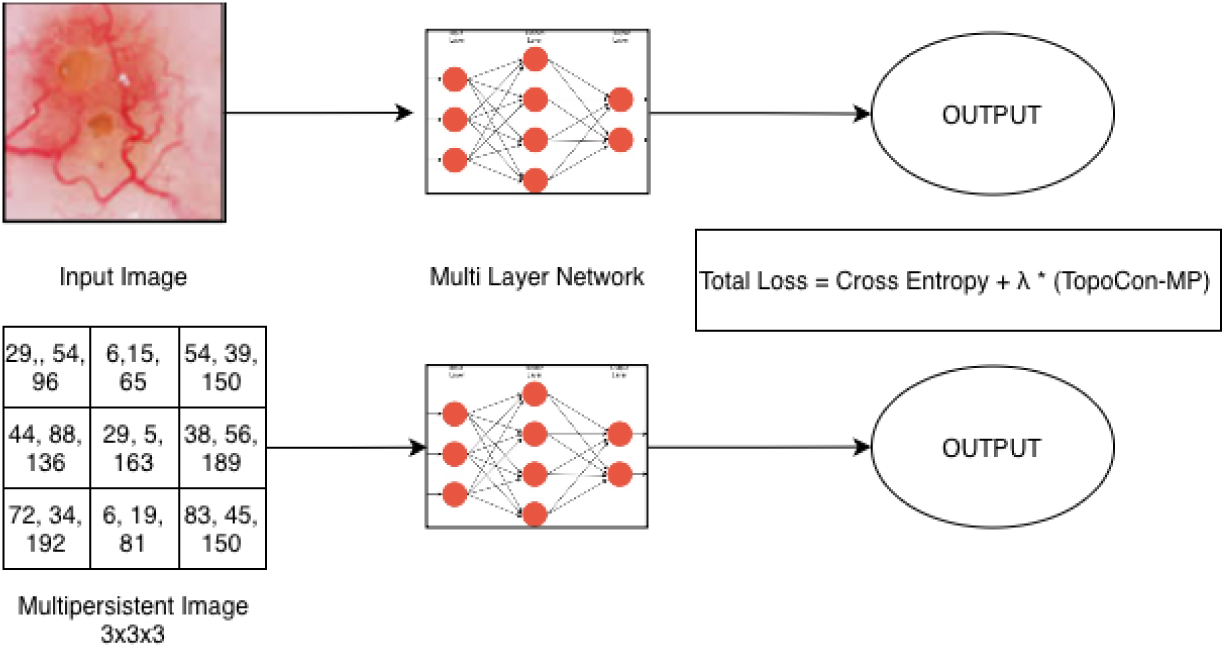
Overview of the topology aware supervised contrastive framework. The raw image and its multipersistence topological image are encoded by separate networks; the resulting embeddings are used both for classification and for a supervised contrastive loss that aligns image and topology representations.

An image encoder *f_θ_*(·) (a pretrained Swin Transformer backbone with a linear head) and a topology encoder *g_ϕ_*(·) (an MLP on Ψ(*I*)) produce latent embeddings *z_I_* = *f_θ_*(*I*)*, z_T_* = *g_ϕ_*(Ψ(*I*)). These embeddings are concatenated and fed to a classifier for the main lesion classification task. In parallel, they are mapped through projection heads and used in a supervised contrastive loss: samples that share the same class label, whether they come from the image branch or the topology branch, are treated as positives, and samples from different classes are treated as negatives (following the formulation of Khosla et al.).

The final training objective combines cross entropy and supervised contrastive losses,

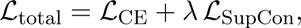

where *λ* balances discriminative and alignment terms. This objective aligns image and topology embeddings in a class consistent manner while preserving classification performance. By leveraging topological representations as label preserving views instead of relying solely on random augmentations, TopoCon-MP provides contrastive supervision that is better tailored to limited data and anatomy sensitive medical imaging settings.

## 4. Experiments

### 4.1. Setup

#### Datasets

We evaluate our framework on three publicly available dermoscopic image datasets: DermaMNIST, MILK-10K, and PAD-UFES-20. Together they cover a range of lesion types, acquisition devices, and class distributions. All images are dermoscopic RGB scans annotated by dermatology experts and are resized to a fixed resolution before training.

DermaMNIST (Yang et al., 2023) is derived from the HAM10000 dataset (Tschandl et al., 2018), part of the ISIC 2018 challenge (Codella et al., 2019). It contains 10,015 dermoscopic images from seven diagnostic categories and serves as a standard benchmark for skin lesion classification.

**Table 1:** Summary of datasets.

| Dataset | # Images | # Classes |
| --- | --- | --- |
| DermaMNIST | 10,015 | 7 |
| MILK-10K | 5,240 | 11 |
| PAD-UFES-20 | 2,298 | 6 |

MILK-10K (Philipp et al., 2025) is a collection of approximately 10,000 dermoscopic and clinical images across eleven classes. To keep the setting consistent across datasets, we use only the dermoscopic subset (5,240 images) in our experiments.

PAD-UFES-20 (Pacheco et al., 2020) comprises 2,298 smartphone-acquired dermoscopic images labeled by pathologists into six diagnostic categories, introducing realistic variability in acquisition conditions and illumination.

For each dataset we perform an 80:10:10 split into training, validation, and test sets (grouped by patient identifier where available to avoid leakage). We first train and evaluate models on each dataset independently. Furthermore, to assess **cross-dataset generalization**, we also conduct a transfer experiment: models are trained on the five lesion classes common to DermaMNIST and the dermoscopic subset of MILK-10K, and evaluated on the corresponding subset of PAD-UFES-20.

#### Preprocessing and Topological Features

All dermoscopic images are resized to 224× 224 pixels to ensure uniform spatial resolution across datasets. Standard normalization is applied to the RGB channels before feature extraction.

##### Single Persistence Topological Descriptors

For the single persistent homology setting, we compute Betti features using 50 filtration thresholds (*n bins* = 50). For each image, this produces 50-dimensional vectors for Betti-0, Betti-1, and the number of activated pixels, computed independently across the RGB and grayscale channels. This results in a total of 4 (channels) × 3 (feature types) × 50 = 600 topological features per image.

##### MultiPersistence Topological Descriptors

To capture richer structural interactions, we further compute cubical multi-parameter persistence using bifiltration over the red and green channels with 20 thresholds each. This yields a 20 × 20 bifiltration grid for each image, producing a 2D topological map with three channels corresponding to Betti-0, Betti-1, and activated pixels calculated during bifiltration. When flattened, this representation provides 20 × 20 × 3 = 1200 multi-parameter topological features per image.

These topological features are first evaluated independently using XGBoost classifiers to assess their discriminative capability. Subsequently, the multi-parameter topological maps are integrated into the **TopoCon-MP** framework, where they are combined with Swin Transformer embeddings for supervised contrastive learning and classification.

#### Hyperparameters

We used the Adam optimizer with a learning rate of 1e^−4^, batch size of 64, and cross-entropy loss with class weights for all experiments. All images were resized to 224 × 224 and normalized with the ImageNet mean and standard deviation. Baseline CNN and ViT models were trained for 15 epochs without augmentation, using ImageNet-pretrained backbones with frozen weights while keeping BatchNorm statistics and Dropout layers active. For the transfer learning setup (training on MILK-10K and DermaMNIST, testing on PAD-UFES-20), we used identical hyperparameters and applied early stopping based on validation macro-F1 (patience = 3).

**Table 2:**
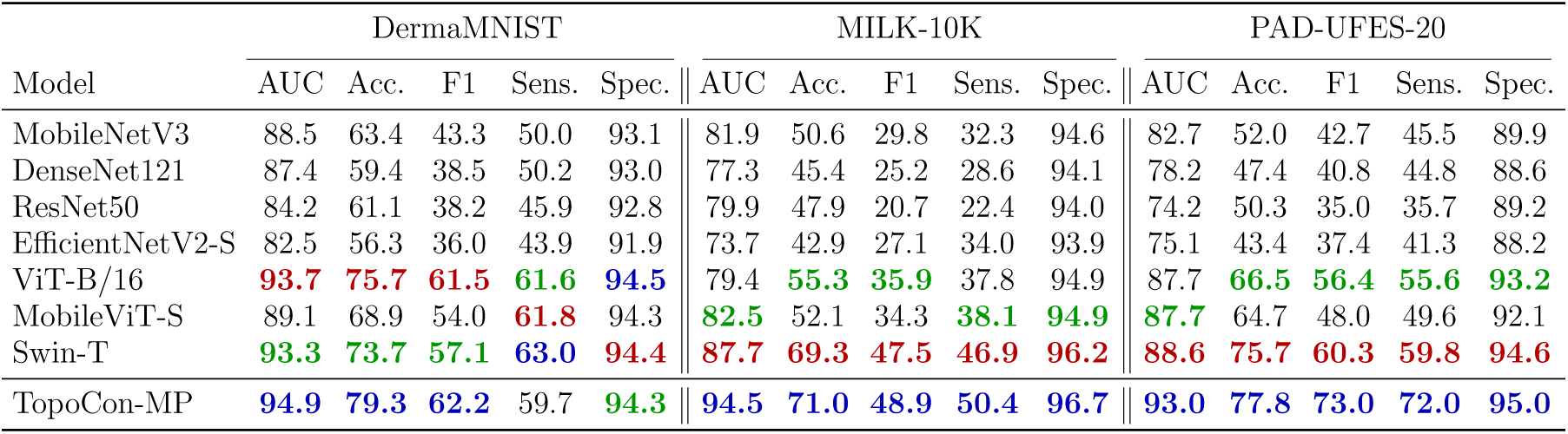
Baseline comparison of 2D CNN and Transformer models with our proposed TopoCon-MP model. The **best**, **second**, and **third** performances across mod-els are highlighted.

Our contrastive fusion model (TopoCon-MP) was trained with the AdamW optimizer (weight decay of 1e^−2^) for 30 epochs under cosine-annealing scheduling. The model fused Swin-T image embeddings with 3 × 20 × 20 topological maps derived from 1200-dimensional multi-persistence vectors. Each vector was normalized in 400-dimensional blocks and re-shaped to a 3 × 20 × 20 tensor. The fusion module consisted of LayerNorm, a linear layer, ReLU activation, and a dropout rate of 0.3. The projection head for contrastive learning was a two-layer MLP with dimensions 768 → 256 → 64, LayerNorm and ReLU activations, mapping fused embeddings to a 64-dimensional latent space. We used a supervised contrastive loss with *λ* = 0.1 and temperature *τ* = 0.07, combined with cross-entropy loss for classification. All models used automatic mixed precision (AMP), TensorFloat-32 (TF32), and gradient clipping at 1.0. The best model was selected based on the highest validation AUC.

### 4.2. Results

#### Baselines

We compare our approach against widely used 2D convolutional and transformer-based architectures. For convolutional baselines, we include MobileNetV3-Large-100 (Howard et al., 2019), DenseNet121 (Huang et al., 2017), ResNet50 (He et al., 2016), and EfficientNetV2-S (Tan and Le, 2021), representing compact, densely connected, residual, and compound-scaled network families respectively. For transformer-style baselines, we evaluate ViT-B/16 (Dosovitskiy et al., 2021), MobileViT-S (Mehta and Rastegari, 2022), and Swin-T (Liu et al., 2021), covering both pure and hybrid vision transformer designs. All models use ImageNet pretrained weights and are fine tuned on the dermoscopic datasets with identical optimization and augmentation settings.

## Discussion

The following table summarizes the quantitative performance of all baseline models and our proposed contrastive fusion method with multi-parameter topology (Contrastive-MP) across the three dermoscopic datasets.

**Table 3:** Baseline comparison of 2D CNN and Transformer models with our proposed TopoCon-MP model. The **best**, **second**, and **third** performances across models are highlighted.

| Model | DermaMNIST |  |  |  |  | MILK-10K |  |  |  |  | PAD-UFES-20 |  |  |  |  |
| --- | --- | --- | --- | --- | --- | --- | --- | --- | --- | --- | --- | --- | --- | --- | --- |
|  | AUC | Acc. | F1 | Sens. | Spec. | AUC | Acc. | F1 | Sens. | Spec. | AUC | Acc. | F1 | Sens. | Spec. |
| MobileNetV3 | 88.5 | 63.4 | 43.3 | 50.0 | 93.1 | 81.9 | 50.6 | 29.8 | 32.3 | 94.6 | 82.7 | 52.0 | 42.7 | 45.5 | 89.9 |
| DenseNet121 | 87.4 | 59.4 | 38.5 | 50.2 | 93.0 | 77.3 | 45.4 | 25.2 | 28.6 | 94.1 | 78.2 | 47.4 | 40.8 | 44.8 | 88.6 |
| ResNet50 | 84.2 | 61.1 | 38.2 | 45.9 | 92.8 | 79.9 | 47.9 | 20.7 | 22.4 | 94.0 | 74.2 | 50.3 | 35.0 | 35.7 | 89.2 |
| EfficientNetV2-S | 82.5 | 56.3 | 36.0 | 43.9 | 91.9 | 73.7 | 42.9 | 27.1 | 34.0 | 93.9 | 75.1 | 43.4 | 37.4 | 41.3 | 88.2 |
| ViT-B/16 | **93.7** | **75.7** | **61.5** | **61.6** | **94.5** | **79.4** | **55.3** | **35.9** | **37.8** | **94.9** | **87.7** | **66.5** | **56.4** | **55.6** | **93.2** |
| MobileViT-S | 89.1 | 68.9 | 54.0 | **61.8** | **94.3** | **82.5** | 52.1 | 34.3 | **38.1** | **94.9** | **87.7** | 64.7 | 48.0 | 49.6 | 92.1 |
| Swin-T | **93.3** | **73.7** | **57.1** | **63.0** | **94.4** | **87.7** | **69.3** | **47.5** | **46.9** | **96.2** | **88.6** | **75.7** | **60.3** | **59.8** | **94.6** |
| TopoCon-MP | **94.9** | **79.3** | **62.2** | 59.7 | **94.3** | **94.5** | **71.0** | **48.9** | **50.4** | **96.7** | **93.0** | **77.8** | **73.0** | **72.0** | **95.0** |

Across all three datasets, our proposed Contrastive-MP model achieves the highest AUC compared to all CNN and transformer baselines. This consistent gain demonstrates the effectiveness of integrating multi-parameter topological features for improved class separation and robustness.

To further evaluate cross-domain generalization, we conduct a transfer learning experiment where models are trained on the common five classes shared between DermaMNIST and MILK-10K, and tested on PAD-UFES-20. Table 4 presents the corresponding results under this domain shift setting.

**Table 4:** Cross-dataset transfer results from models trained on DermaMNIST and the dermoscopic subset of MILK-10K and evaluated on PAD-UFES-20. For each metric, the **best**, **second, and third performances across models are high-lighted.**

| Model | AUC | Acc | F1 | B.Acc | Sens | Spec |
| --- | --- | --- | --- | --- | --- | --- |
| MobileNetV3 | 66.2 | 39.3 | 30.9 | 32.5 | 32.5 | 82.8 |
| DenseNet121 | 66.6 | 36.6 | 30.3 | 31.6 | 31.6 | 81.7 |
| ResNet50 | 64.0 | 39.8 | 20.8 | 24.7 | 24.7 | 80.6 |
| EfficientNetV2-S | 62.4 | 34.9 | 27.2 | 29.3 | 29.3 | 82.4 |
| ViT-B/16 | **78.9** | **51.3** | **45.5** | **46.8** | **46.8** | **86.2** |
| MobileViT-S | 73.8 | 41.1 | 33.9 | 38.8 | 38.8 | 83.8 |
| Swin-T | **78.7** | **47.6** | **42.4** | **46.3** | **46.3** | **86.1** |
| TopoCon-MP | **78.5** | **50.3** | **36.7** | **64.2** | **42.9** | **85.5** |

### Ablation Studies

We conduct two ablation studies to disentangle the contributions of the topological representation and the fusion strategy. First, in Table 5 we compare single parameter (SP) cubical persistence on grayscale and on all RGB channels with our multiparameter red plus green (MP RG) encoding, using the same XGBoost classifier. On DermaMNIST, SP on all channels achieves the highest AUC, reflecting that a simple multi channel filtration already captures much of the topology in this relatively clean benchmark. On MILK-10K and especially on the more heterogeneous PAD UFES 20 dataset, however, MP RG matches or exceeds SP in AUC and yields clear gains in accuracy, F1, and balanced accuracy, indicating that multipersistence becomes more beneficial under varied acquisition conditions. Second, Table 6 evaluates different learners on the same 3 × 20 × 20 multipersistence tensors. XGBoost provides a strong non deep baseline, while a Swin backbone trained only on the topological image struggles and is unstable on MILK 10K and PAD UFES 20. In contrast, our full TopoCon MP model (MP+SupCon) that jointly trains the image and topology encoders with supervised contrastive alignment delivers consistent and often large improvements across all datasets and metrics, showing that both the MP features and the topology aware contrastive fusion are essential for the final performance.

**Table 5:**
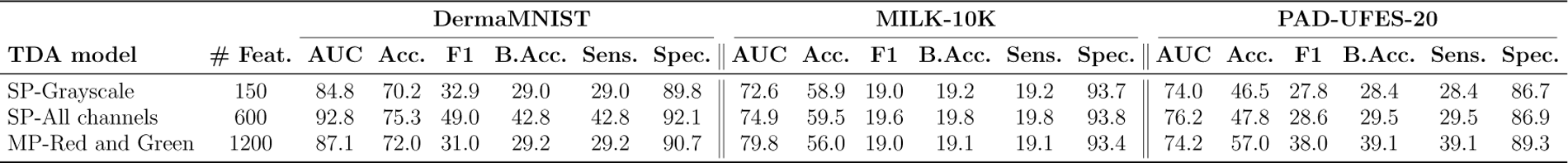
Topological Feature Ablation. The results of our ablation study of XGBoost model on topological features across different channels and datasets

**Table 6:**
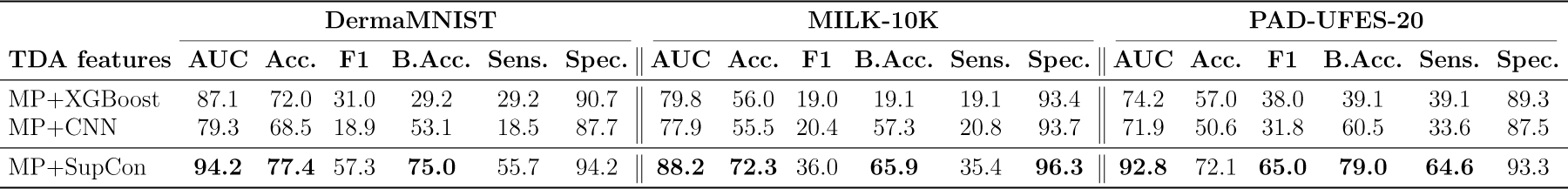
ML Ablation. The performances of different ML models utilizing our 3×20×20 multipersistence outputs.

### Visualization

We also investigate what the multipersistence descriptors learn by visualizing them on MILK-10K. Appendix A.1 presents two complementary views: Figure 7 shows mean (*β*_0_) and (*β*_1_) Betti curves with confidence bands across color channels, while Figure 8 displays classwise median red–green Betti tensors and activated-pixel maps. Together, these plots reveal consistent, class-specific patterns in multiscale topology (for example, broader peaks and shifted hotspots for melanoma and keratinocytic lesions), providing qualitative evidence that our multipersistence features capture clinically meaningful lesion structure rather than arbitrary handcrafted cues. In Figure 5, we also include DermaMNIST red-channel (*β*_0_) curves, which exhibit analogous class-specific differences in the evolution of connected components.

**Figure 5:**
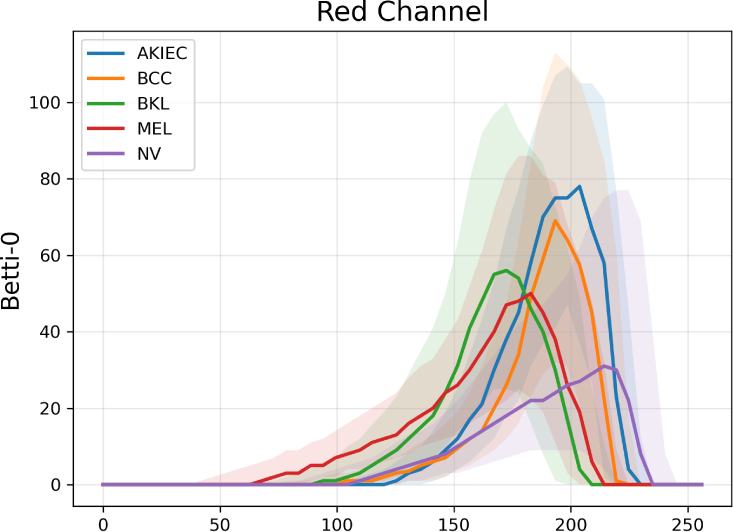
DermaMNIST Betti-0 curves (red channel). Mean *β*_0_ curves with 40% confidence bands for DermaMNIST lesion classes (AKIEC, BCC, BKL, MEL, NV) as a function of red-channel intensity threshold, showing class-specific patterns in the evolution of connected components.

## 5. Conclusion

In summary, we introduced cubical multiparameter persistence for dermoscopic image analysis and showed that topology provides complementary signal to modern vision backbones in skin cancer classification. Across multiple public datasets, multipersistence alone was competitive and, when aligned with a Vision Transformer via supervised contrastive learning, consistently outperformed strong pretrained CNN and ViT baselines in accuracy and AUC. Ablations confirmed that multiparameter topology yields richer cues than single-parameter cubical persistence and that contrastive alignment is critical for effective fusion. Beyond performance, the topological channels offered interpretable patterns that co-localized with clinically meaningful structures, supporting more transparent decision making. Limitations include the need to choose filtration parameters and grids, the computational overhead of multipersistence, and evaluation focused primarily on dermoscopy. Future work will explore adaptive and differentiable multiparameter filtrations, tighter end-to-end training with vision models, uncertainty quantification, and extensions to segmentation, multimodal fusion, and prospective clinical studies.

## Data Availability

All data produced are available online at https://medmnist.com/ and
https://challenge.isic-archive.com/landing/milk10k/

https://challenge.isic-archive.com/landing/milk10k

## Acknowledgments

This work was partially supported by National Science Foundation under grants DMS-2220613, and DMS-2229417. The authors acknowledge the Texas Advanced Computing Center (TACC) at UT Austin for providing computational resources that have contributed to the research results reported within this paper.

## Appendix

## Appendix A. Multiparameter Persistence

Multiparameter persistence has attracted growing interest for its potential to enrich standard persistent homology. In principle, a multidimensional filtration with several parameters should yield richer topological summaries for machine learning than a one-parameter filtration. However, key technical obstacles have limited its practical impact.

In single-parameter persistence, the threshold space {*α_i_*} is totally ordered, so each topological feature in the filtration {Δ*_i_*} has well defined birth and death times. This makes it possible to decompose the associated persistence module 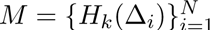 into a multiset of intervals (barcodes) via a structure theorem (Botnan and Lesnick, 2022), which underlies persistence diagrams. For two or more parameters, the threshold set {(*α_i_, β_j_*)} is only partially ordered. Birth and death times are no longer uniquely defined, the one dimensional decomposition theorem does not extend (Botnan and Lesnick, 2022), and barcode representations typically fail to exist or are difficult to describe in a finite way. As a result, a direct barcode style generalization of single-parameter persistence is usually not available, and the classification and invariants of multiparameter modules remain an active area of research in commutative algebra (Eisenbud, 2013).

Despite these challenges, several slicing based methods have been proposed to make use of multiparameter filtrations (Lesnick, 2015; Carrière and Blumberg, 2020; Botnan and Lesnick, 2022). These approaches analyze one dimensional slices of the multiparameter grid, compute standard persistence diagrams along each slice, and then aggregate the resulting diagrams into vectorized summaries. While effective in some settings, they face two main limitations: the summaries can depend strongly on the choice of slicing directions, and compressing information from many diagrams into a low dimensional representation may introduce substantial information loss; see (Botnan and Lesnick, 2022) for a detailed overview.

In this work we adopt a different strategy that avoids slicing. We work directly with the Betti numbers on the grid: for each grid point and each homological dimension *k*, we record the rank of *H_k_* and collect these ranks into Betti tensors. This can be viewed as evaluating the Hilbert function of the underlying multiparameter module on a fixed finite grid. The resulting Betti tensors provide a simple, grid aligned summary that is easy to compute and to feed into neural networks, and they have been empirically effective in several imaging applications (Qaiser et al., 2019; Yadav et al., 2023; Du et al., 2022; Ali et al., 2023).

### Other bifiltration examples

A well known limitation of grayscale sublevel filtrations is their inability to encode the *size* of topological features; they only reflect differences in function values between the birth and death of a feature. For example, consider a grayscale image where all pixels have intensity 0 except for a single central pixel with intensity 255. The resulting persistence diagram contains a single long bar [0, 255), even though the corresponding hole has diameter 1. Conversely, a binary image 𝒳_100_ might contain a large hole of diameter 20 whose pixels have intensities in [101, 105], so the hole is completely filled by 𝒳_105_. Despite the dramatic change in geometric size, the grayscale sublevel filtration produces only a short bar (100, 105), encoding the contrast but not the spatial scale of the hole.

**Figure 6:**
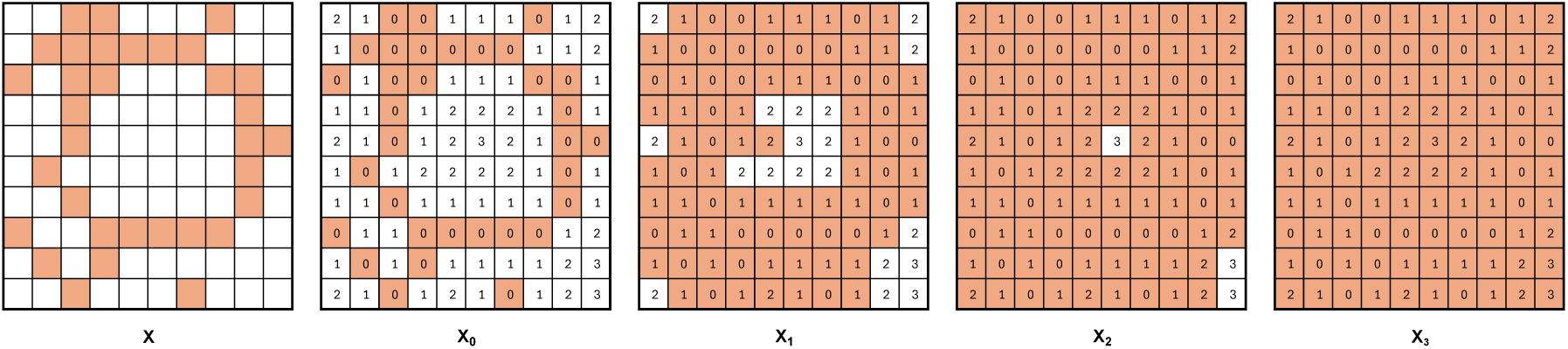
Erosion filtration. For a given binary image 𝒳, we first define the erosion function (shown in 𝒳_0_). We then obtain a filtration of binary images 𝒳_0_ ⊂ 𝒳_1_ ⊂ · · · ⊂ 𝒳_3_ by activating pixels that reach the threshold value.

In other words, while persistent homology identifies which topological features appear in a filtration, standard sublevel filtrations do not, by themselves, capture their geometric size. To address this, alternative filtrations such as *erosion, dilation*, and *signed-distance based filtrations* have been proposed (Garin and Tauzin, 2019). These constructions explicitly incorporate scale information and thus complement grayscale sublevel filtrations. In particular, one can combine a grayscale or color channel with an erosion or distance based filtration to obtain meaningful multiparameter persistence signatures for images.

## A.1. Visualizations and Interpretability of Topological Descriptors

To better understand what information our multipersistence descriptors capture, we visualize classwise Betti curves and median multipersistence heatmaps for the MILK-10K dataset.

### Betti curves across color channels

Figure 7 shows the mean Betti curves with 40% confidence bands for five lesion classes (BCC, NV, BKL, MEL, AKIEC) across red, green, blue, and grayscale filtrations. The top row plots *β*_0_ (number of connected components) as a function of the intensity threshold and the bottom row plots *β*_1_ (number of holes). The solid (top) and dashed (bottom) lines denote the classwise means, while the shaded regions mark the central 40% of subjects for each class.

**Figure 7:**
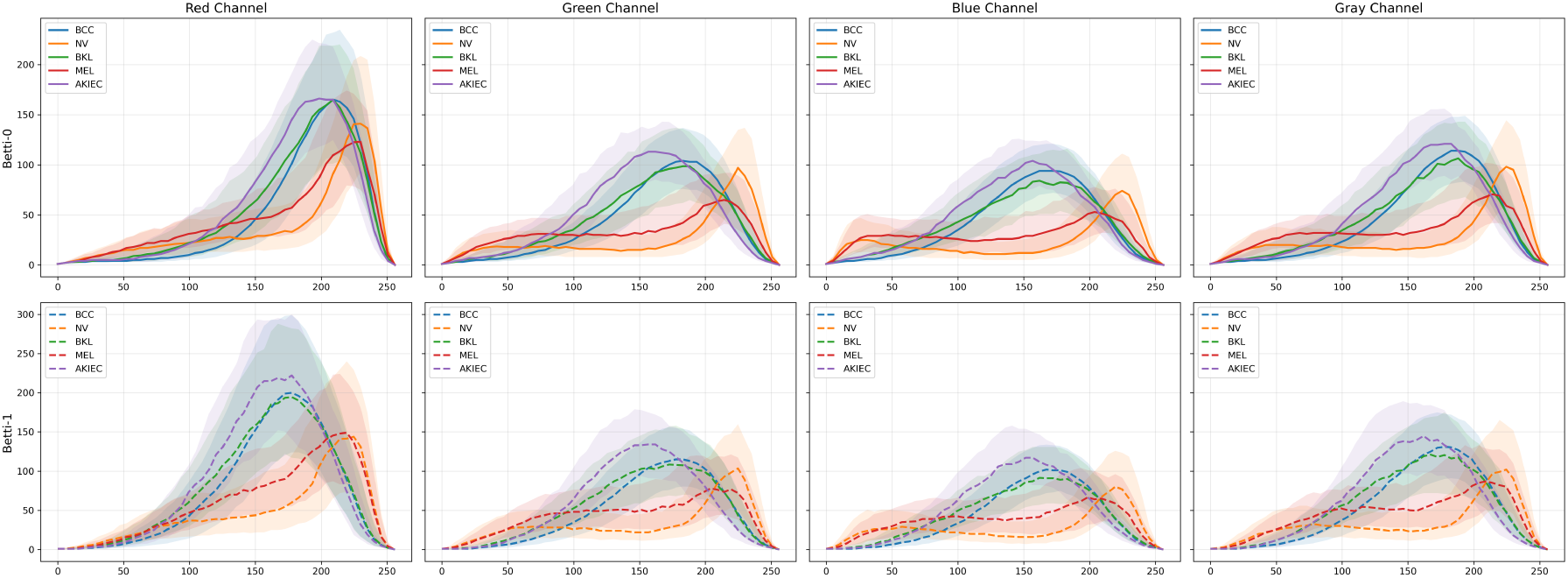
Betti curves for MILK-10K. Mean Betti curves with 40% confidence bands for five MILK-10K lesion classes (BCC, NV, BKL, MEL, AKIEC) across color channels. Columns correspond to red, green, blue, and grayscale intensity filtrations. The top row shows *β*_0_ (number of connected components) and the bottom row shows *β*_1_ (number of holes) as functions of the threshold value. Solid (top) and dashed (bottom) lines indicate classwise means, and shaded regions mark the central 40% of subjects per class, revealing systematic differences in multiscale topology between lesion types.

Several consistent patterns emerge. First, BKL and AKIEC lesions tend to exhibit higher and broader *β*_0_ and *β*_1_ peaks, reflecting a larger number of small islands and holes across intermediate thresholds. This is compatible with their irregular, mottled pigmentation patterns in dermoscopy. In contrast, NV lesions show lower-amplitude curves and narrower peaks, consistent with more homogeneous, compact nevi. MEL curves typically peak at slightly darker thresholds than NV, suggesting richer structure in darker pigment regions, which is in line with the irregular networks and focal globules often seen in melanoma. Across channels, the red and grayscale filtrations yield the strongest separation between classes, which motivated our choice of red–green bifiltrations for the main multipersistence pipeline.

### Multipersistence heatmaps

While Betti curves summarize topology along a single intensity axis, our model uses bifiltrations over red and green channels. Figure 8 displays classwise median multipersistence heatmaps on the 20 × 20 red–green grid. Each row corresponds to one lesion class, and the three columns show *β*_0_, *β*_1_, and activated-pixel counts, respectively. Color encodes the median value at each grid point, so bright regions indicate parameter ranges where many connected components, holes, or pixels are present.

**Figure 8:**
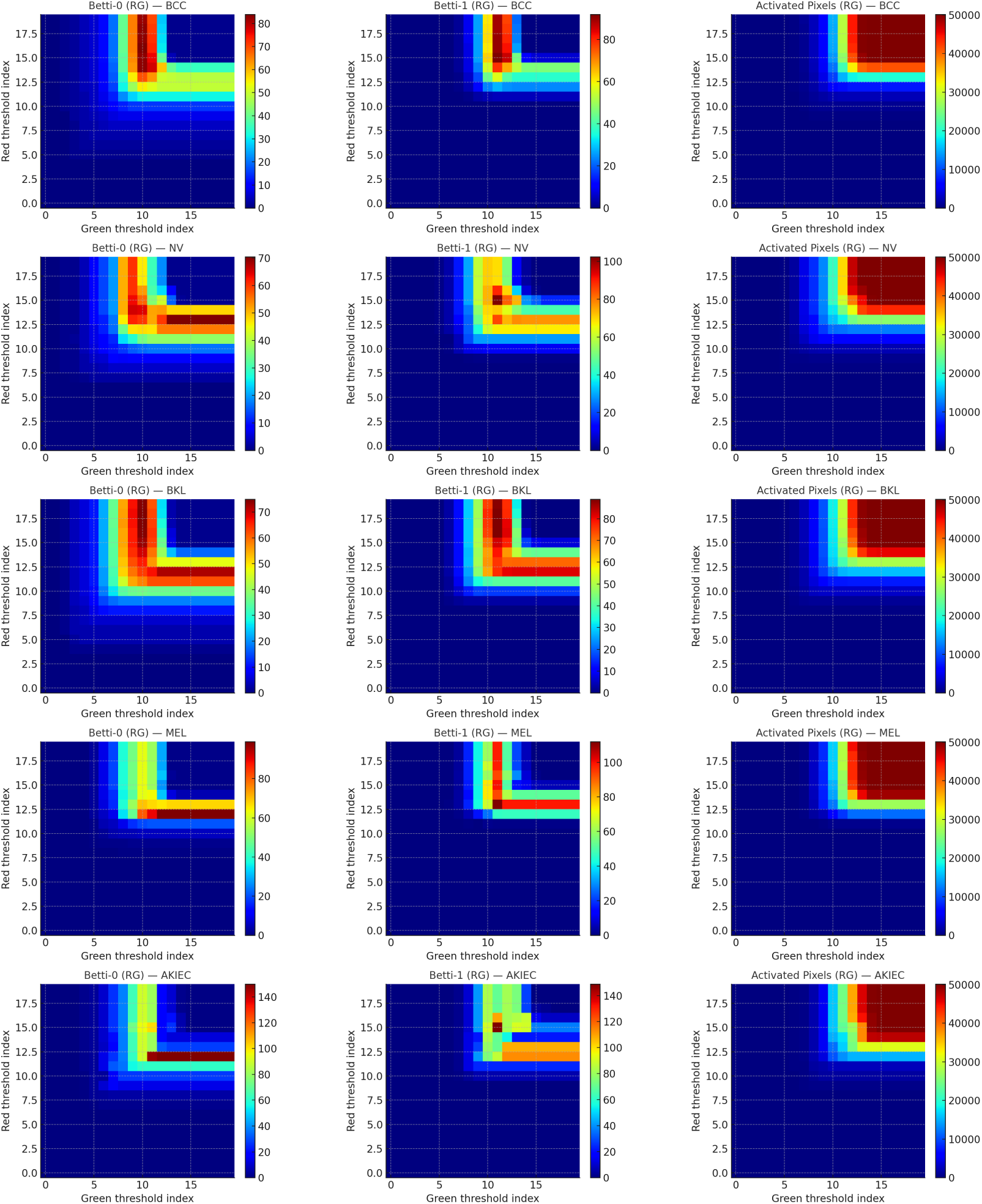
Multipersistence heatmaps for MILK-10K. Classwise median multiparameter descriptors computed on the red–green bifiltration grid (20 × 20 thresholds). Rows correspond to lesion classes (BCC, NV, BKL, MEL, AKIEC) and columns show *β*_0_, *β*_1_, and activated-pixel counts, respectively. Color encodes the median value at each grid point, highlighting distinct patterns of connected components, holes, and overall lesion occupancy across classes in the multipersistence representation.

These heatmaps reveal complementary structure that is not visible from single-channel curves alone. For example, NV lesions concentrate most of their *β*_0_ and *β*_1_ mass in a relatively compact block of lighter thresholds, indicating uniform pigmentation with limited cross-channel variation. BKL and AKIEC classes show broader, more diffuse high-intensity regions that extend toward darker red and green levels, consistent with heterogeneous pigmentation and scattered foci. MEL lesions exhibit a shift of the *β*_1_ hotspot toward darker-red / mid-green thresholds, suggesting complex hole patterns in specific color combinations that align with irregular pigment networks and streaks. The activated-pixel maps further highlight differences in overall lesion occupancy in the red–green plane, which our model uses jointly with *β*_0_ and *β*_1_.

Overall, these visualizations support the view that multipersistence encodes class-specific, multiscale topology that is coherent with known dermoscopic morphologies. While we do not claim these descriptors are directly diagnostic on their own, they offer an interpretable intermediate representation: the regions of the red–green grid where our Betti tensors are most active correspond to characteristic patterns of lesion fragmentation and hole formation that our TopoCon-MP model can exploit during training.

